# Using HiFi Long-Read Whole Genome Sequencing To Enhance Diagnosis In Patients With Subfertility And/Or Recurrent Pregnancy Loss

**DOI:** 10.64898/2026.05.01.26352136

**Authors:** Teo Jing Xian, Chanatjit Cheawsamoot, Donghyeon Kim, Jasmine Goh Chew Yin, Sylvia Kam, Shermaine Chan Soak Mun, Yang Liying, Liu Shuling, Chua Khi Pin, Wilson Cheng, Gwo-Chin Ma, Ting-Yu Chang, Keh-Ming Wu, Yi-Shing Lin, Eun Jeong Yu, Yeseul Kim, Moon-Woo Seong, Punkavee Tuntiviriyapun, Paweena Thuwanut, Chanakarn Suebthawinkul, Chalurmpon Srichomthong, Wanna Chetruengchai, Songphon Kanlayaprasit, Rutairat Wongong, Jonas Korlach, Jee-Soo Lee, Ming Chen, Sohyun Hwang, Weng Khong Lim, Vorasuk Shotelersuk, Saumya Shekhar Jamuar

## Abstract

Subfertility and recurrent pregnancy loss (RPL) affect a significant proportion of couples worldwide. Genetic causes can be seen in up to 30% of these individuals but require multiple genetic tests, which often impede a comprehensive work up. Newer genomic technologies, such as PacBio HiFi long read sequencing (LRS) can detect most subclasses of variations (such as structural rearrangement, monogenic disorders) through one single test.

In this multicenter study, we enrolled couples with unexplained subfertility and/or RPL and performed HiFi LRS to determine the underlying genetic etiology. Participants were recruited using a standardized inclusion/ exclusion criteria to rule out other known causes of subfertility and/or RPL. 96 individuals were recruited across the 5 sites. Average age of participants was 36 years (range 30-46 years). Among the 84 individuals who completed sequencing, 4.8% were identified with a likely genetic diagnosis and variants of uncertain significance were identified in another 14.2% of individuals. One individual was identified with an ACMG secondary finding, and while multiple carriers for recessive genetic disorders were identified, none of the couples were identified to be at increased risk.

This study highlights the utility of performing genomic sequencing in couples with unexplained subfertility and/or RPL, with 1 in 10 couples harboring a clinically significant variant. In addition, use of HiFi LRS allowed for characterization of different subclasses of genomic variations through a single test. Future studies, including exploring the cost effectiveness and resource utilization of LRS as first line test, will help in optimizing care for such couples.

**TWEETABLE STATEMENT:** A single long-read genome sequencing test can consolidate multiple genetic investigations and uncover clinically relevant causes in couples with unexplained subfertility and recurrent pregnancy loss.

**AT A GLANCE:**

A. Why was this study conducted?
  - Many couples with subfertility and recurrent pregnancy loss remain undiagnosed after multiple conventional genetic tests
  - Existing workflows require sequential testing and may miss complex genomic variants
B. What are the key findings?
  - Long-read genome sequencing identified clinically relevant variants in ∼1 in 10 couples with unexplained subfertility or recurrent pregnancy loss
  - A single assay enabled detection of multiple variant types, including structural and sequence variants
C. What does this study add to what is already known?
  - Demonstrates feasibility of a unified genomic testing approach in a real-world multicenter cohort
  - Supports a potential shift from fragmented testing toward a single comprehensive genomic workflow

## Introduction

Subfertility is estimated to affect one in six people worldwide, while 1-2% of women suffer from recurrent pregnancy losses (RPL)^1^. Together, these conditions result in significant psychological and financial impact on affected couples and pose considerable management and counselling challenges for clinicians. Parental chromosomal anomalies such as translocations and inversions have been reported to occur in 3-5% of couples with RPL as compared to 0.7% of the general population, while genetic causes are estimated to contribute to approximately 30% of male and female subfertility cases^2,3,4^. Newer evidence also suggests that complex chromosomal rearrangements (CCRs) and submicroscopic chromosomal abnormalities are associated with infertility and RPL^5^. Identification of these abnormalities and accurate characterization of apparently balanced chromosome rearrangements in individuals with subfertility and RPL can therefore provide important information for counselling on reproductive risk and tailoring assisted reproductive treatments.

Conventional karyotyping is typically performed in patients with subfertility and RPL but can only identify large structural rearrangements (>5 Mb). Molecular techniques such as chromosomal microarray (CMA) and whole-exome sequencing (WES) cannot identify balanced translocations, inversions and CCRs. Whole genome sequencing (WGS) is a short-read sequencing technology that is superior in detecting single nucleotide variations (SNVs) and indels but limited in the ability to detect copy number variants (CNVs) and not able to detect methylation or repeat expansions^6^.

Newer genomic technologies, such as long-read sequencing (LRS) by Pacific Biosciences can not only detect SNVs, indels, but also CNVs, SVs, methylation and repeat expansion^7^. LRS may therefore be used to not only more precisely identify CCRs but also elucidate information on changes in submicroscopic copy numbers and provide information on the parental origin of new chromosomal breakpoints. This data can be used to tailor assisted reproductive treatment, for example by allowing for preimplantation genetic testing for structural rearrangements (PGT-SR), to assist in embryo selection and potentially improve conception and live birth rates.

The effectiveness of using long-read sequencing as the first line diagnostic test for patients with subfertility and recurrent miscarriages has not been well studied. Herein, we aimed to study the impact of the diagnostic journey of patients with unexplained subfertility and/or RPL with the use of PacBio HiFi long read whole genome sequencing. We hypothesized that LRS will be able to identify all the relevant types of variants in patients with subfertility and RPLs through a single test, thereby reducing the diagnostic journey for the patient and potentially impacting reproductive outcomes by enabling personalized assisted reproductive treatment.

## Methods and Approach

### HiFi Solves Subfertility Consortium in Asia-Pacific

The HiFi Solves Sub-fertility Consortium is an innovative collaboration involving five leading centers across the Asia-Pacific region-Maternal Child Health Research Institute (MCHRI), including KK Women’s and Children’s Hospital, Singapore (KKH) and Singapore General Hospital (SGH), Singapore; Center of Excellence for Medical Genomics, Chulalongkorn University, Thailand (CU); Changhua Christian Hospital Medical Center, Taiwan (CCHMC); Seoul National University College of Medicine, South Korea (SNUCM); and CHA Bundang Medical Center, CHA University School of Medicine, South Korea (CUSM) (Figure 1)^8^. The consortium was established to understand the use of PacBio HiFi long-read sequencing to enhance the diagnosis care of individuals with subfertility and/or RPL.

**Figure 1.**
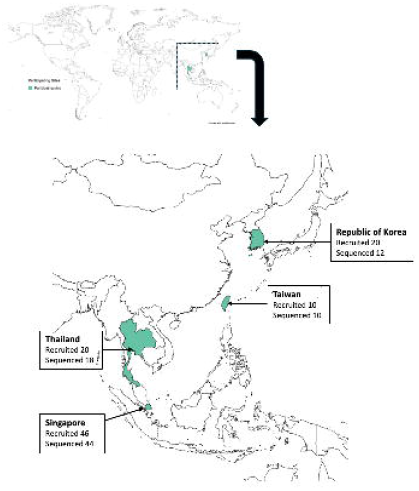
Schematic showing the participating centers and the recruitment across each site

### Patient recruitment

Patients were recruited at each of the 5 sites through a local IRB approved project (MCHRI-CIRB 2019/2243 (approved 19 Aug 2014), CU-RA-MF-14/68 (approved 28 Feb 2025), CCHMC-240513 (approved 14 Jul 2024), SNUCM-H-2503-113-1623 (approved 25 Apr 2025), CUSM-GCI 2024-11-004-002 (approved 16 Jan 2025)). Informed consent was obtained from the participant and partner, where available.

### Inclusion/ exclusion criteria

#### Inclusion criteria

- Patients/couples with 3 or more pregnancy losses before the age of 38 years and normal karyotype
- Unexplained fertility of 1 or more years before the age of 38 years for females and 40 years for males

#### Exclusion criteria

- Known chromosomal abnormalities
- Known past medical/surgical history that can explain RPL (e.g. autoimmune disease, thrombophilia, uterine anomalies, etc.) or subfertility (e.g. abnormal semen analysis, blocked tubes, anovulation, etc.)

### Clinical data

Data was collected as per standardized data collection form (Appendix 1). Briefly, this included demographic data of the patient and the partner, family history, medical history and results of past investigations, including genetic testing and imaging.

### HiFi Sequencing and bioinformatics analysis

Genomic DNA extracted from 96 participants were used. DNA was sheared to ∼15 kb fragments, and DNA quality and quantity were assessed by Qubit and pulse-field gel electrophoresis. Around 6μg of DNA was used for SMRTbell® Prep Kit 3.0 library preparation. Libraries were sequenced on the Revio (Pacific Biosciences, USA) system using one SMRT Cell per sample. Alignment^9,10,11,12^ and variant discovery were performed using the comprehensive PacBio HiFi-human-WGS-WDL pipeline^13^ (v3.1.0 (KKH), v3.0.1 (CHA) and v2.2.1 (CU)). Full tool versioning is detailed in the tool’s documentation page (https://github.com/PacificBiosciences/HiFi-human-WGS-DL/blob/fb65f901b30c7ae2fda533ef39f28b1019de856c/docs/tools_containers.md). Briefly, HiFi reads were aligned to GRCh38 reference genome using pbmm2 v1.17.0. Single-nucleotide variants (SNVs) and indels were called using DeepVariant v1.9.0, while structural variants (SVs) and copy number variants (CNVs) were identified via Sawfish v2.1.0. Tandem repeat genotyping was performed using TRGT v4.0.0.

To establish haplotype-resolved variant sets, SNVs, indels, and SVs were jointly phased using HiPhase v1.5.0. Additionally, highly homologous genes within segmental duplications (including *SMN1/2, HBA1/2*, and *HBB*) were targeted for specialized genotyping using Paraphase v3.3.4. For epigenetic analysis, 5mC probabilities were summarized at GRCh38 CpG sites using pb-CpG-tools v3.0.0.

### Variant prioritization and validation

SNVs and indels were annotated using VEP release 112. SVs and CNVs were annotated using AnnotSV v3.5. QC filters for variants included: 1) have ≥10× total read depth and 2) show an alternate allele fraction of 20–80% for heterozygous calls. Clinical prioritization of candidate variants was conducted using a strategy based on looking at Phrank scores calculated via the pipeline using patient-specific Human Phenotype Ontology (HPO) terms (Appendix 2) and the genes associated with subfertility/ recurrent pregnancy loss (Appendix 3). For SNVs, additional filters included variant and genotype quality ≥Q20, allele frequency ≤1% in gnomAD v4.1 and in-house local databases (such as Thai Exome cohort (n=7000), Thai Long Read cohort (n=300) for CU samples and CHORUS^14^ for KKH samples), and pathogenicity predictions (REVEL ≥0.644 for missense; SpliceAI ≥0.2 for splicing). SVs and CNVs were further required to involve coding regions and have allele frequency ≤1% in both the Database of Genomic Variants and our in-house cohort. Single nucleotide variants (SNV) and Copy number variants (CNVs) were classified according to ACMG/AMP and Riggs et al^15,16^ guidelines, respectively, and those without prior reports were considered novel.

## Results

### Demographics

A total of 96 individuals were recruited for this study-this included 47 couples and 2 individuals. For the 2 individuals, only the female partner was recruited. Average age of participants was 36 years (range 30-46 years), with individuals of Chinese, Korean and Thai ethnicity representing 32.6%, 23.3% and 20.9% of the cohort. Out of the cohort, 39 couples presented with recurrent pregnancy loss, 20 couples presented with subfertility and 7 couples with infertility (Supp Table 1)

### QC metrics-depth, quality

From the 96 individuals, DNA from 93 samples passed the QC threshold for HiFi LRS. For the 3 failed samples, one was recollected and passed QC threshold, while the other two individuals declined repeat blood draw. Overall, 86 samples were sequenced across four sites (KKH (n=44), CHA (n=12), CU (n=20) and CCHMC (n=10)). However, two samples from the CU site were noted to have a similar variant profile, which upon review was identified due to mislabeling of samples, where the same sample was sequenced twice. The couple declined repeat blood draw and were excluded from subsequent analysis.

Overall, 84 samples were sequenced to a mean depth of 34.6x, with an average yield of 106.8Gb and a read N50 of 16.5kb (Supp table 2). Variant calling identified a mean of 4.1 million SNVs, 855,064 indels, and 38,526 structural variants per individual.

### Bioinformatics analysis linked to primary phenotype

Following multi-step filtering, 44 candidate variants were prioritized in 28 individuals (Supp Table 3). This included 39 SNVs, 3 indels and 2 CNVs. None of the individuals had any pathogenic repeat expansions, including at the Fragile X locus. These variants were further curated using the ACMG/AMP variant curation framework. Where available, medical records were reviewed to correlate genomic findings with clinical phenotype to further refine the curation. Five variants were classified as P/LP, 33 as VUS and 6 as likely benign (Table 1, 2, 3, Supp Table 4).

**Table 1.**
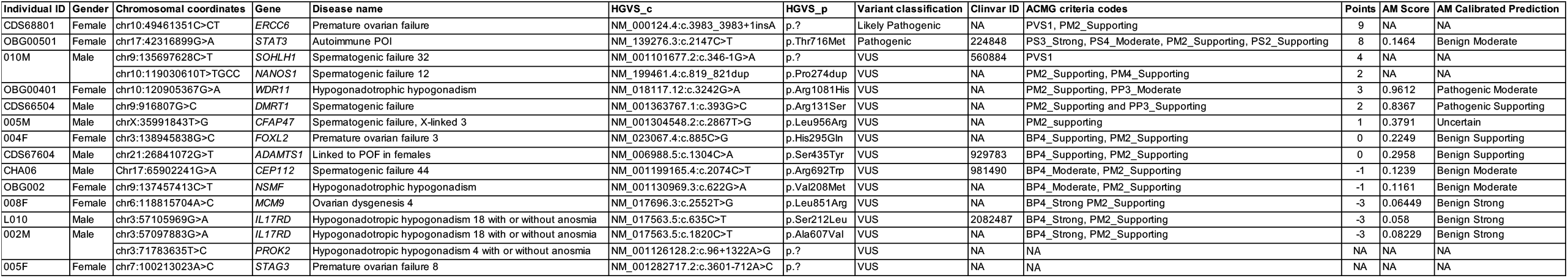
Variant information in genes associated with monoallelic inheritance.

**Table 2.**
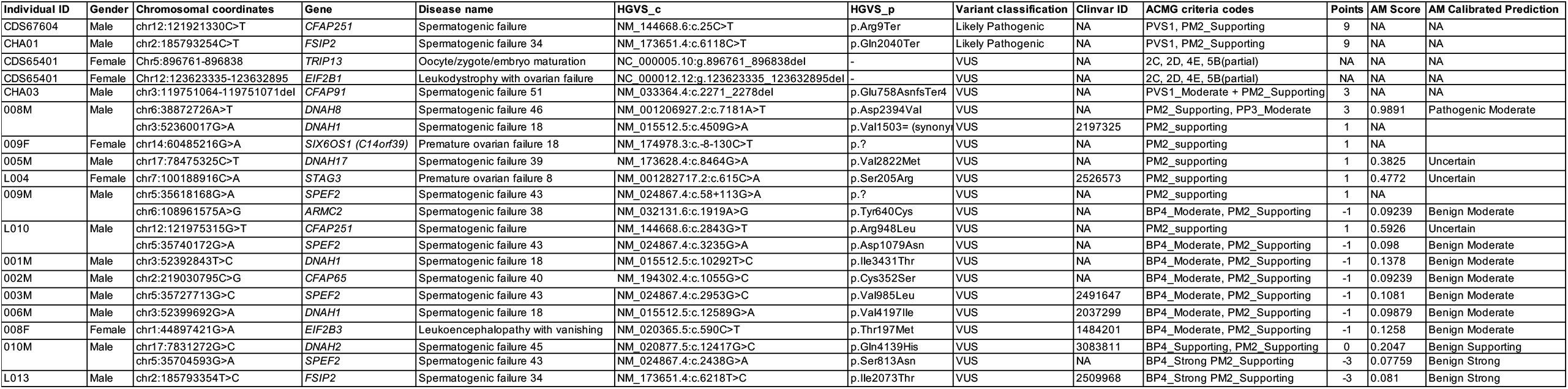
Variant information in genes associated with biallelic inheritance.

**Table 3.** Summary of variants identified across each category-primary phenotype, ACMG findings, carrier status.

|  | Primary finding-<br>monoallelic<br>(n=84) | Primary<br>finding-<br>biallelic (n=84) | ACMG<br>incidental<br>finding (n=74) | Carrier finding<br>(n=74) |
| --- | --- | --- | --- | --- |
| **P/LP** | 2 | 2 | 1 | 35 |
| **VUS ($\geq 3$)** | 2 | 4 | NA | NA |
| **VUS<3** | 12 | 16 | NA | NA |

#### Candidate variants in genes associated with monoallelic inheritance

In genes associated with a dominant inheritance (n=15) or X-linked in males (n=1) and/or where haploinsufficiency or gain of function is a known or possible disease mechanism, 2 variants were classified as P/LP and 14 as VUS giving an overall rate of identifying a candidate variant at 19.0% (Table 1).

Among the two individuals with pathogenic/ likely pathogenic variants, one female (OBG00501) had a recurrent pathogenic variant in *STAT3* associated with autoimmune POI (OMIM# 615952), and one female (CDS68801) had a novel frameshift variant in a gene associated with ovarian dysgenesis (*ERCC6* (OMIM #616946). Two individuals had VUS leaning towards pathogenicity. These included splicing variant in *SOHLH1* (OMIM# 618115), associated with spermatogenic failure, in a Chinese male (010M) with normal karyotype and normospermia, and a novel missense variant in *WDR11* (OMIM# 614858), associated with hypogonadotropic hypogonadism, in a Chinese female (OBG00401) with recurrent pregnancy losses and low level mosaic monosomy X. Based on these findings, the presumed diagnostic yield is 4.8%.

Twelve additional VUS were identified in 11 individuals. These variants were in genes associated with POI (*STAG3* (OMIM# 615723), *ADAMTS1* (OMIM# 605174)); spermatogenic failure (*DMRT1* (OMIM# 602424), *DMRT1* (OMIM# 602424), *NANOS1* (OMIM# 615413), *CEP112* (OMIM# 619044) and *CFAP47* (OMIM# 301059)); and hypogonadotropic hypogonadism (*NSMF* (OMIM# 614838), *IL17RD* (OMIM# 615267) and *PROK2* (OMIM# 610628)). Functionalization of these variants would be required to provide further clarity on the causativeness of these variants but that is beyond the scope of this study.

#### Candidate variants in genes associated with biallelic inheritance

Eighteen individuals had a candidate variant in genes associated with biallelic variant (Table 2). All but four individuals had one variant each, while the remaining four (008M, 009M, 010M and L010) had 2 variants each. Two males had a likely pathogenic variant in genes associated with recessively inherited spermatogenic failure (*CFAP251* (OMIM# 618152) and *FSIP2* (OMIM# 618153)). Sixteen individuals had VUS in genes associated with either spermatogenic failure (n=15) and ovarian failure (n=4). One individual has variant in a gene associated with embryo maturation arrest (*TRIP13* (OMIM# 619011)).

Review of the other allele on the LRS data did not identify any other rare variant, both in the coding as well as non-coding regions, in these individuals suggesting that these individuals were likely to be carriers, rather than affected by these diseases. While less likely, we are unable to rule out a second mosaic variant in the gonadal tissue as that is beyond the scope of this study.

### Secondary findings

Given that HiFi LRS generated data beyond the genes associated with primary phenotype, we reviewed the individuals’ genomic data for additional findings, including those in the ACMG SF list^17^ as well as for recessive genetic diseases, as per Mackenzie Mission list^18^. We included variants that were either classified as pathogenic and/or likely pathogenic in Clinvar, and/or novel loss of function variants in recessive genetic diseases.

We completed this analysis in 74 individuals. Among them, only one (1.4%) had a secondary finding in the *LPL* gene, associated with hypercholesterolemia (OMIM#144250) (Supp Table 5). Twenty-six (35.1%) individuals were identified to be carriers for at least one recessive genetic disease. Except for one couple (CDS723), who were known to be carriers for deletion in *REN*, there were no other couples who were carriers for the same genetic disease (Supp Table 6). One individual (CDS70604) had two variants in *DUOX2* associated with thyroid dyshormonogenesis (OMIM# 607200). However, both variants were on the same allele suggesting that the individual is a carrier, rather than affected with thyroid dyshormonogenesis.

## Discussion

The HiFi Solves Sub-Fertility Consortium was established as a multicenter study aimed to understand the utility of PacBio HiFi LRS in couples with unexplained subfertility/ infertility and/or RPL. In this study, 96 individuals were recruited across four sites in Asia (Singapore, Thailand, Taiwan and Korea) and genomic data was available from 84 individuals. The diagnostic yield for primary phenotype was 4.8%, suggesting that ∼1 in 10 affected couples would directly benefit from genetic testing. Moreover, the disease-causing variants were present in both females and males, highlighting the need to test both partners as part of the diagnostic work up.

LRS provided a few distinct advantages. Beyond making a diagnosis in 4.8% of the cohort, we identified individuals with one pathogenic variant in recessively inherited genes, LRS enabled us to rule out any other rare coding and/or noncoding variants in the second allele, providing greater certainty with regards to the significance of these variants. Next, for the remaining couples with no candidate variant, comprehensively ruling out known genetic causes is also valuable information allowing them to focus on other possible (especially non-genetic) causes of subfertility/ RPL. Lastly, 26 individuals were identified to be carriers of a recessive genetic disease, and 1 individual has a secondary finding (in *LPL* gene), consistent with literature^22,23^, highlighting the advantage of performing broader genomic wide testing in investigating such couples as it provides information regarding their personal and reproductive risk beyond the primary phenotype. Moreover, one individual had two disease-causing variants in the same gene. Long read enabled phasing showing that the two variants were on the same allele, hence, reducing the need for additional testing in this individual.

Traditional testing for genetic etiology in affected couples involves performing karyotype, chromosomal microarray analysis, Fragile X analysis, followed by either panel or exome/genome sequencing. However, LRS can analyze each of the subclasses of genetic variants through one single test^3^. Not only does LRS provide information about the different variant subclass, but it also provides information relating to secondary findings and carrier status, which can be helpful for couples in their family planning journey.

### Limitations

As with any genomic tests, we identified multiple VUS in genes associated with either ovarian or spermatogenic failure, or hypogonadotropic hypogonadism. Additional hormonal profiling, semen analysis and/or functionalization of these variants would help to clarify the significance of these variants and may potentially further increase the diagnostic yield^19,20,21^ but is beyond the scope of the study.

While LRS can detect multiple subclasses, the ability to identify balanced translocations, a frequent cause of subfertility and recurrent pregnancy loss, remains a challenge due to paucity of LRS data to identify the true positives. This is expected to improve over time as more normative data is generated. However, in the interim, couples need the additional karyotype, which was performed in our cohort, to rule out balanced translocation.

Finally, the cost of LRS is higher compared to the traditional genetic tests, However, as the cost of LRS continues to drop, future studies may be able to demonstrate the cost effectiveness and reduced healthcare utilization when performing LRS as the first line test for such couples.

## Conclusion

Despite the small sample size, we demonstrate the potential utility of using LRS in couples with unexplained subfertility/infertility and/or recurrent pregnancy loss, with ∼1 in 10 couples harboring a clinically relevant genomic variant. These findings support the potential role of long-read genome sequencing as an early comprehensive test in the diagnostic evaluation of couples with unexplained subfertility and recurrent pregnancy loss, and future studies with larger sample sizes in conjunction with declining costs and improve bioinformatics analytical pipelines may highlight the role of LRS as the first line test for such couples.

## Acknowledgements

We thank the families for participating in this study, study and all the staff at the Reproductive Biology Unit, King Chulalongkorn Memorial Hospital, Faculty of Medicine, Chulalongkorn University, Thailand, for their assistance and contribution during participant recruitment and sample collection.

We thank the additional members from Seoul National University Hospital for their contributions to patient recruitment, data collection, and clinical support, including Hyesu Lee, Seung Won Chae, Hobin Sung, Sung Im Cho, JiYeon Han, Joo Won Jang, Hoyeon Lee, Hoon Kim, and Seung-Yup Ku.

We also thank all participating sites and clinical teams across the HiFi Solves Sub-Fertility Consortium for their contributions.

## Data availability statement

The data used in this study are not publicly available due to privacy and legal restrictions. Patients IDs have been deidentified and are not known to anyone outside the research group.

## Consent

Informed consent was obtained from all participants (MCHRI-CIRB 2019/2243 (approved 19 Aug 2014), CU-RA-MF-14/68 (approved 28 Feb 2025), CCHMC-240513 (approved 14 Jul 2024), SNUCM-H-2503-113-1623 (approved 25 Apr 2025), CUSM-GCI 2024-11-004-002 (approved 16 Jan 2025)).

## Supplementary Data

### Text

**Appendix 1:** Participant recruitment form

**Appendix 2:** List of HPO terms

**Appendix 3:** List of genes associated with phenotype

### Tables

**Supplementary Table 1:** Demographic information of participants

**Supplementary Table 2:** QC metrics across the sites

**Supplementary Table 3:** Candidate variants identified in the individuals

**Supplementary Table 4:** List of variants classified as benign

**Supplementary Table 5:** List of clinically relevant variants in ACMG SF list

**Supplementary Table 6:** List of clinically relevant variants in MM gene list

## References

1. World Health Organization. Infertility prevalence estimates, 1990–2021. Geneva: WHO; 2023.

2. El Hachem H, Crepaux V, May-Panloup P, Descamps P, Legendre G, Bouet PE. Recurrent pregnancy loss: current perspectives. Int J Womens Health. 2017;9:331–345.

3. Veltman JA, Tüttelmann F. Why geneticists should care about male infertility. Nat Rev Genet. 2024. doi:10.1038/s41576-024-00773-3.

4. Krausz C, Riera-Escamilla A. Genetics of male infertility. Nat Rev Urol. 2018;15(6):369–384. doi:10.1038/s41585-018-0003-3.

5. Liu L, Kang K, Wang H, Sun L, et al. Accurate detection of multiple chromosome rearrangements and copy number variations by PacBio sequencing in complex chromosomal abnormality. Prenat Diagn. 2026. doi:10.1002/pd.70120.

6. Kernohan KD, Boycott KM. The expanding diagnostic toolbox for rare genetic diseases. Nat Rev Genet. 2024;25(6):401–415. doi:10.1038/s41576-023-00683-w.

7. Marx V. Method of the year: long-read sequencing. Nat Methods. 2023;20(1):6–11. doi:10.1038/s41592-022-01730-w.

8. Pacific Biosciences. PacBio announces the HiFi Solves Sub-fertility Consortium in Asia-Pacific [Internet]. Available from: https://www.pacb.com/press_releases/pacbio-announces-the-hifi-solves-sub-fertility-consortium-in-asia-pacific/

9. McLaren W, Gil L, Hunt SE, Riat HS, Ritchie GR, Thormann A, et al. The Ensembl Variant Effect Predictor. Genome Biol. 2016;17:122.

10. Geoffroy V, Herenger Y, Kress A, Stoetzel C, Piton A, Dollfus H, et al. AnnotSV: an integrated tool for structural variations annotation. Bioinformatics. 2018;34(20):3572–3574.

11. Jagadeesh KA, Wenger AM, Berger MJ, Guturu H, Stenson PD, Cooper DN, et al. Phrank measures phenotype sets similarity to improve Mendelian disease diagnosis. Genet Med. 2019;21(2):464–470.

12. Thormann A, Halachev M, McLaren W, Moore D, Svinti V, Campbell A, et al. Flexible and scalable diagnostic filtering of genomic variants using G2P with Ensembl VEP. Nat Commun. 2019;10:2373.

13. Lake JA, Concepcion G, Ward H, Rowell W, Wenger AM, Fang K, et al. HiFi-human-WGS-WDL (v3.1.0): PacBio HiFi human whole genome sequencing analysis pipeline [Internet]. Zenodo; 2025. Available from: https://github.com/PacificBiosciences/HiFi-human-WGS-WDL/releases/tag/v3.1.0

14. Genome Institute of Singapore, Agency for Science, Technology and Research (ASTAR). National Precision Medicine (NPM) programme (SG10K_Health) [Internet]. Singapore: ASTAR; [cited 2026 Apr 16]. Available from: https://www.a-star.edu.sg/gis/our-science/precision-medicine-and-population-genomics/npm

15. Riggs ER, Andersen EF, Cherry AM, Kantarci S, Kearney H, et al. Technical standards for the interpretation and reporting of constitutional copy-number variants: a joint consensus recommendation of the American College of Medical Genetics and Genomics (ACMG) and the Clinical Genome Resource (ClinGen). Genet Med. 2020;22(2):245–257. doi:10.1038/s41436-019-0686-8. :contentReference[oaicite:0]{index=0}

16. Richards S, Aziz N, Bale S, Bick D, Das S, Gastier-Foster J, Grody WW, Hegde M, Lyon E, Spector E, Voelkerding K, Rehm HL; ACMG Laboratory Quality Assurance Committee. Standards and guidelines for the interpretation of sequence variants: a joint consensus recommendation of the American College of Medical Genetics and Genomics and the Association for Molecular Pathology. Genet Med. 2015;17(5):405–424. doi:10.1038/gim.2015.30.

17. Miller DT, Lee K, Abul-Husn NS, Amendola LM, Brothers K, Chung WK, Gollob MH, Gordon AS, Harrison SM, Hershberger RE, Klein TE, Richards CS, Stewart DR, Martin CL; ACMG Secondary Findings Working Group. Electronic address. ACMG SF v3.2 list for reporting of secondary findings in clinical exome and genome sequencing: A policy statement of the American College of Medical Genetics and Genomics (ACMG). Genet Med. 2023 Aug;25(8):100866. doi: 10.1016/j.gim.2023.100866.

18. Delatycki MB, Laing NG, Aitken M, et al. Preconception carrier screening for genetic conditions: The Mackenzie’s Mission. Med J Aust. 2020;213(11):506–509. doi:10.5694/mja2.50853.

19. Stallmeyer B, Tüttelmann F, Riera-Escamilla A, et al. How exome sequencing improves the diagnostics and understanding of male infertility. Andrology. 2024. doi:10.1111/andr.13728.

20. Nagirnaja L, Aston KI, Conrad DF, et al. Diverse monogenic subforms of human spermatogenic failure. Nat Commun. 2022;13:7953. doi:10.1038/s41467-022-35661-z.

21. Bazrgar M. Genetics of female infertility. Front Endocrinol (Lausanne). 2023;14:1244567. doi:10.3389/fendo.2023.1244567.

22. SG10K_Health Consortium, Chan SH, Bylstra Y, Teo JX, Kuan JL, Bertin N, et al. Analysis of clinically relevant variants from ancestrally diverse Asian genomes. Nat Commun. 2022;13:6694. doi:10.1038/s41467-022-34116-9.

23. Machini K, Ceyhan-Birsoy O, Azzariti DR, et al. Analyzing and reanalyzing the genome: findings from the MedSeq Project. Am J Hum Genet. 2019;105(1):177–188. doi:10.1016/j.ajhg.2019.05.017.

