## Supplemental Text 1 for "Using HiFi Long-Read Whole Genome Sequencing To Enhance Diagnosis In Patients With Subfertility And/Or Recurrent Pregnancy Loss"

Supplementary Text

Appendix 1: Data collection sheet

**Recurrent pregnancy loss/ Subfertility Research**

Family Study ID:

DEMOGRAPHICS

*Wife’s details*

·	Name:

·	Date of birth (DD/MM/YY):

·	Ethnicity:

·	Personal medical history: _ Yes	_ No

If yes, please specify:

*Husband’s details*

·	Name:

·	Date of birth (DD/MM/YY)

·	Ethnicity:

·	Personal medical history: _ Yes	_ No

If yes, please specify:

*Eligibility criteria*

·	Unexplained recurrent pregnancy loss: _ Yes	_ No

o	If yes, please specify number of pregnancy losses:

·	Unexplained subfertility: _ Yes	_ No

o	If yes, please specify:

*Past investigations*

·	US pelvis: _ Normal	_ Abnormal	_ Not done

o	If abnormal, please specify:

·	Autoimmune screen: _ Normal	_ Abnormal	_ Not done

o	If abnormal, please specify:

·	Karyotype: _ Normal	_ Abnormal	_ Not done

o	If abnormal, please specify:

·	Fragile X testing (for wife): _ Normal	_ Abnormal	_ Not done

o	If abnormal, please specify:

·	Chromosome Y deletion (for husband): _ Normal	_ Abnormal	_ Not done

o	If abnormal, please specify:

·	Cystic fibrosis testing: _ Normal	_ Abnormal	_ Not done

o	If abnormal, please specify:

*Family history*

Recurrent miscarriages: _ Yes	_ No

If yes, please specify relationship

Stillborn: _ Yes	_ No

If yes, please specify:

Congenital disorders: _ Yes	_ No

If yes, please specify:

Learning disability: _ Yes	_ No

If yes, please specify:

Consanguinity: _ Yes	_ No

If yes, please specify:

**NOTES:**

Age for females: 21 years (inclusive) to 37 years (inclusive);

**Not pregnant**

Age for males: 21 years (inclusive) to 40 years (inclusive)

|  | Inclusion Criteria | Exclusion Criteria |
| --- | --- | --- |
| Unexplained recurrent pregnancy loss | - 3 or more pregnancy losses **before age of 38**  - Normal karyotype  - Recruit as a couple (i.e. 1 couple = 2 participants)    *For females recruited under this group, upper age limit does not apply. | - Known chromosomal abnormalities.  - Known past medical/surgical history that can explain RPL (i.e. autoimmune disease, uterine anomalies) – this criteria to be defined by the OBGYN specialist |
| Unexplained subfertility | - Subfertility of 1 or more years |  |

*Name and date of birth to be replaced by Study ID (Family Study ID PLUS M for male, F for female) and age (in years) when sharing datasheet with collaborators
