## Supplemental Text 2 for "Using HiFi Long-Read Whole Genome Sequencing To Enhance Diagnosis In Patients With Subfertility And/Or Recurrent Pregnancy Loss"

Appendix 2

List of HPO terms

- Decreased fertility (HP:0000144)
- Female infertility (HP:0000737)
- Male infertility (HP:0003251)
- Abnormality of the ovary (HP:0000137)
- Anovulation or primary amenorrhea (HP:0000786)
- Recurrent pregnancy loss (HP:0001191)
- Habitual abortion and Spontaneous abortion (HP:0001281)
- Recurrent spontaneous abortion (HP:0001191)
