## Supplemental Text 3 for "Using HiFi Long-Read Whole Genome Sequencing To Enhance Diagnosis In Patients With Subfertility And/Or Recurrent Pregnancy Loss"

Appendix 3

Gene panels

Female subfertility

*AIRE, AKR1C4, ANOS1, AR, ARL6, ARX, ATRX, AXL, BBS1, BBS10, BBS12, BBS2, BBS4, BBS5, BBS7, BBS9, BMP15, CBX2, CCDC141, CHD7, CLPP, CYP11A1, CYP17A1, CYP19A1, DHH, DMRT1, DMRT2, DUSP6, EIF2B1, EIF2B2, EIF2B3, EIF2B4, EIF2B5, FEZF1, FGF17, FGF8, FGFR1, FGFR2, FIGLA, FLRT3, FOXL2, FSHB, FSHR, GALT, GNRH1, GNRHR, HESX1, HFE, HFM1, HOXA13, HS6ST1, HSD17B3, IL17RD, KISS1, KISS1R, LEP, LEPR, LHB, LHCGR, LHX3, LHX4, LMNA, MAMLD1, MAP3K1, MCM8, MCM9, MKKS, NOBOX, NR0B1, NR3C1, NR5A1, NSMF, NUP107, PADI6, PCSK1, POR, PRLR, PROK2, PROKR2, PROP1, PSMC3IP, RSPO1, SEMA3A, SEMA3E, SOHLH1, SOX10, SOX2, SOX3, SOX9, SPRY4, SRA1, SRD5A2, SRY, STAG3, STAR, SYCE1, TAC3, TACR3, TRIM32, TTC8, WDR11, WNT4, WT1, WWOX, ZP1*

Male subfertility

*ACTL9, ADGRG2, AIRE, AKR1C4, AMH, AMHR2, ANOS1, AR, ARL6, ARMC2, ARX, ATRX, AURKC, AXL, BBS1, BBS10, BBS12, BBS2, BBS4, BBS5, BBS7, BBS9, BRDT, C14orf39, CATSPER1, CBX2, CCDC141, CEP112, CFAP251, CFAP43, CFAP44, CFAP47, CFAP58, CFAP65, CFAP69, CFAP70, CFAP91, CFTR, CHD7, CYP11A1, CYP17A1, CYP19A1, DHH, DMRT2, DNAH1, DNAH10, DNAH17, DNAH2, DNAH6, DNAH8, DUSP6, DZIP1, FANCM, FEZF1, FGF17, FGF8, FGFR1, FGFR2, FLRT3, FOXL2, FSHB, FSIP2, GALNTL5, GATA4, GNRH1, GNRHR, HESX1, HFE, HS6ST1, HSD17B3, IL17RD, INSL3, KISS1, KISS1R, KLHL10, LEP, LEPR, LHB, LHCGR, LHX3, LHX4, M1AP, MAMLD1, MAP3K1, MKKS, NANOS1, NPAS2, NR0B1, NR5A1, NSMF, PCSK1, PICK1, PLCZ1, PMFBP1, PNLDC1, POR, PPP2R3C, PROK2, PROKR2, PROP1, QRICH2, RSPO1, SEMA3A, SEMA3E, SEPTIN12, SLC26A8, SOHLH1, SOX10, SOX2, SOX3, SOX9, SPATA16, SPEF2, SPRY4, SRA1, SRD5A2, SRY, STAR, SUN5, SYCE1, SYCP2, SYCP3, TAC3, TACR3, TAF4B, TEX11, TEX14, TEX15, TRIM32, TTC21A, TTC29, TTC8, WDR11, WNT4, WT1, WWOX, ZMYND15*
