## Supplemental Table 1 for "Using HiFi Long-Read Whole Genome Sequencing To Enhance Diagnosis In Patients With Subfertility And/Or Recurrent Pregnancy Loss"

| Sample ID | Gender | Ethnicity | Relationship | Phenotype |
|---|---|---|---|---|
| *Maternal and Child Health Research Institute, Singapore* | | | | |
| CDS65401 | Female | Chinese | Self | Unexplained subfertility<br>Karyotype: 46,XX |
| CDS65404 | Male | Chinese | Husband | Unexplained subfertility<br>Karyotype: 46,XY |
| CDS66101 | Female | Chinese | Self | Unexplained subfertility<br>Karyotype: 46,XX |
| CDS67601 | Female | Indonesian | Self | Recurrent Pregnancy Loss<br>Karyotype: Not done |
| CDS67604 | Male | Indonesian | Husband | Recurrent pregnancy loss<br>Karyotype: Not done |
| OBG00101 | Female | Malay | Self | Recurrent pregnancy loss X3.<br>Benign inversion carrier |
| OBG00104 | Male | Javanese | Husband | Normal |
| CDS66601 | Female | Chinese | Self | Unexplained subfertility<br>Karyotype: Not done |
| CDS66604 | Male | Chinese | Husband | Unexplained subfertility<br>Karyotype: Not done |
| CDS66501 | Female | Chinese | Self | Unexplained subfertility<br>Karyotype: Not done |
| CDS66504 | Male | Chinese | Husband | Unexplained subfertility<br>Karyotype: Not done |
| OBG00301 | Female | Indian | Self | Recurrent pregnancy loss X6; karyotype normal |
| OBG00304 | Male | Indian | Husband | karyotype Normal;Sperm DNA fragmentation 30 % |
| OBG00201 | Female | Indian | Self | Recurrent pregnancy loss X3. 46,XX i.e. normal |
| OBG00204 | Male | Indian | Husband | karyotype is 46,XY,(16qh+) which is a normal variant |
| CDS67201 | Female | Chinese | Self | Recurrent pregnancy loss<br>Karyotype (TMC): 46,XX |
| CDS67204 | Male | Chinese | Husband | Recurrent pregnancy loss<br>Karyotype (TMC): 46,XY |
| CDS67801 | Female | Chinese | Self | Recurrent pregnancy loss<br>Karyotype: 46,XX |
| CDS67804 | Male | Burmese | Husband | Recurrent pregnancy loss<br>Karyotype: 46,XY |
| OBG00401 | Female | Chinese | Self | Recurrent pregnancy loss X5, Culture indicates a female karyotype with mosaic 45,X/46,XX.Low level 45,X/46,XX mosaicism in phenotypically normal |
| CDS68501 | Female | Chinese | Self | Recurrent pregnancy loss<br>Karyotype: 46,XX |
| CDS68504 | Male | Chinese | Husband | Recurrent pregnancy loss<br>Karyotype: 46,XY |
| CDS68701 | Female | Chinese | Self | Recurrent pregnancy loss<br>Karyotype: 46,XX |
| CDS68704 | Male | Chinese | Husband | Recurrent pregnancy loss<br>Karyotype: 46,XY |
| CDS68801 | Female | Chinese | Self | Recurrent pregnancy loss<br>Karyotype (TMC): 46,XX |
| CDS68804 | Male | Chinese | Husband | Recurrent pregnancy loss<br>Karyotype (TMC): 46,XY |
| CDS68901 | Female | Chinese | Self | Recurrent pregnancy loss<br>Karyotype: 46,XX |
| CDS68904 | Male | Chinese | Husband | Recurrent pregnancy loss<br>Karyotype: 46,XY |
| CDS69001 | Female | Chinese | Self | Unexplained subfertility<br>Karyotype: not done |
| CDS69004 | Male | Chinese | Husband | Unexplained subfertility<br>Karyotype: not done |
| CDS69301 | Female | Caucasian | Self | Unexplained subfertility<br>Karyotype: 46,XX |
| CDS69304 | Male | Malay | Husband | Unexplained subfertility<br>Karyotype: 46,XY |
| CDS69801 | Female | Chinese | Self | Unexplained subfertility<br>Karyotype: not done |
| CDS69804 | Male | Chinese | Husband | Husband of CDS69801. Unexplained subfertility |
| OBG00501 | Female | Malay | Self | Recurrent pregnancy loss X5,<br>Karyotype: Culture indicates a female karyotype with no apparent chromosomal abnormalities, |
| OBG00504 | Male | Others | Husband | male karyotype with no apparent chr abnormalities *husband's sample failed QC and declined redraw |
| OBG00601 | Female | Malay | Self | Recurrent pregnancy loss X3, one Child(CS 32 weeks) has ASD and speech delay,<br>Karyotype: Culture indicates a female karyotype with no apparent chromosomal abnormalities |
| OBG00604 | Male | Malay | Husband | Karyotype: Culture indicates a male karyotype with no apparent chromosomal abnormalities |
| OBG00701 | Female | Chinese | Self | Recurrent pregnancy loss X3 |
| OBG00704 | Male | Chinese | Husband | sperm normal |
| CDS70601 | Female | Chinese | Self | Recurrent pregnancy loss |
| CDS70604 | Male | Chinese | Husband | Wife (CDS70604) with recurrent pregnancy loss |

| | | | | |
|---|---|---|---|---|
| CDS72302 | Female | Caucasian | Self | Recurrent pregnancy loss<br>G6P1<br>- Spontaneous pregnancy losss in 2015 and 2016. Karyotype for second pregnancy loss: 46,XX<br>- Third pregnancy: elective LSCS for polyhydramnios. Healthy baby girl delivered.<br>- Fourth pregnancy: FTS, harmony both are low risk for aneuploidies. No obvious fetal anomalies at 12 weeks scan.<br>16 weeks scan: no fetal heart noted, overlapping fetal skull bones and very little amniotic fluid noted.<br>CMA on POC: homozygous, likely pathogenic, deletion of Exons 1-3 of REN gene (biparental inheritance)<br><br>Couple went for IVF+PGT for REN deletion.<br>Had 7 embryos - 4 normal, 2 affected, 1 ?<br>Had two failed IVF+PGT transfer<br>Two successful IVF+PGT transfer<br>- fifth pregnancy: early pregnancy loss at 9 weeks<br>- sixth pregnancy: IUD at 17 weeks. Scan at 16 weeks normal. FetalSeq: no known pathogenic copy number gain/loss in fetus. XX sex chromosomes. |
| CDS72303 | Male | Caucasian | Husband | Recurrent pregnancy loss |
| CDS73001 | Female | Eurasian | Self | Subfertility |
| CDS73004 | Male | Indian | Husband | Unexplained subfertility |
| *CHA University School of Medicine, South Korea* | | | | |
| CHA01 | female | Korean | Self | Subfertility/Recurrent pregnancy loss |
| CHA01 | male | Korean | Husband | |
| CHA02 | female | Korean | Self | Subfertility |
| CHA02 | male | Korean | Husband | |
| CHA03 | female | Korean | Self | Subfertility |
| CHA03 | male | Korean | Husband | |
| CHA04 | female | Korean | Self | Subfertility |
| CHA04 | male | Korean | Husband | |
| CHA05 | female | Korean | Self | Unexplained infertility |
| CHA05 | male | Korean | Husband | |
| CHA06 | female | Korean | Self | Unexplained infertility |
| CHA06 | male | Korean | Husband | |
| CHA07 | female | Korean | Self | Unexplained infertility |
| CHA07 | male | Korean | Husband | |
| CHA08 | female | Korean | Self | Subfertility |
| CHA08 | male | Korean | Husband | |
| CHA09 | female | Korean | Self | Unexplained infertility |
| CHA09 | male | Korean | Husband | |
| CHA10 | female | Korean | Self | Unexplained infertility |
| CHA10 | male | Korean | Husband | |
| *Chulalongkorn University, Thailand* | | | | |
| 001F | Female | Thai | Self | Recurrent Pregnancy Loss: Previous history with natural spontaneous abortion and fail Intrauteine insemination (x 12) |
| 001M | Male | Thai | Husband | . |
| 002F | Female | Thai | Self | Recurrent Pregnancy Loss |
| 002M | Male | Thai | Husband | . |
| 003F | Female | Thai | Self | Recurrent Pregnancy Loss |
| 003M | Male | Thai | Husband | Normospermia |
| 004F | Female | Thai | Self | Recurrent Pregnancy Loss: Spontaneous abortion (x 2: G1 and G2) |
| 004M | Male | Chinese | Husband | Asthenoteratozoospermia |
| 005F | Female | Thai | Self | Recurrent Pregnancy Loss: Previous history with fail embryo transfer (x 4) and natural spontaneous abortion (x 2) |
| 005M | Male | Thai | Husband | . |
| 006F | Female | Thai | Self | Recurrent Pregnancy Loss: Natutal spontaneous abortion (x 3) |
| 006M | Male | Thai | Husband | Normospermaia |
| 007F | Female | Thai | Self | Recurrent Pregnancy Loss: Natutal spontaneous abortion (x 2) and fail frozen embryo transfer (x 1) |
| 007M | Male | Thai | Husband | *sample mislabelled |
| 008F | Female | Thai | Self | Recurrent Pregnancy Loss: Natutal spontaneous abortion (x 1 ), fail intrauterine insemination (x1) and fail frozen embryo transfer (x1) |
| 008M | Male | Thai | Husband | Normal karyotype |
| 009F | Female | Thai | Self | Recurrent Pregnancy Loss |
| 009M | Male | Thai | Husband | . |
| 0010F | Female | Thai | Self | Recurrent Pregnancy Loss: Natutal spontaneous abortion (x 2 ) |
| 0010M | Male | Cninese | Husband | Normal karyotype and norrmospermia |
| Changhua Christian Hospital, Taiwan | | | | |
| L002 | Female | Taiwanese | Self | Recurrent Pregnancy Loss (x2)<br>One pregnancy with an abnormal AF karyotype: 47,XY,+mar(14) dn |
| L003 | Male | Taiwanese | Husband | |

| | | | | |
|---|---|---|---|---|
| L004 | Female | Taiwanese | Self | Infertility<br>Karyotype: 46,XX,r(21)(p13q22.3)dn[21]/45,XX,-21[5]/46,XX,dic r(21;21)(p13q22.3;p13q22.3)dn[2]<br>aCGH: arr[GRCh37] 21q22.3(46761632_48067924)×1 |
| L005 | Male | Taiwanese | Husband | |
| L006 | Female | Taiwanese | Self | Recurrent Pregnancy Loss (x2)<br>Karyotype: 46,XX |
| L007 | Male | Taiwanese | Husband | Karyotype: 46,XY,fra(16)(q22)[4]/46,XY,del(16)(q22){3}/46,XY[18] |
| L009 | Female | Taiwanese | Self | Recurrent Pregnancy Loss (x6) |
| L010 | Male | Taiwanese | Husband | |
| L012 | Female | Chinese | Self | Infertility(10 years)<br>External carrier screening: couple with VUS variants in GCDH gene<br>Desires frozen embryos |
| L013 | Male | Taiwanese | Husband | |
