## Supplemental Table 5 for "Using HiFi Long-Read Whole Genome Sequencing To Enhance Diagnosis In Patients With Subfertility And/Or Recurrent Pregnancy Loss"

| Individual ID | Gender | Ethnicity | Gene | Disease name | Chromosomal coordinates | HGVS_c | HGVS_p | Clinvar Class | Clinvar ID |
|---|---|---|---|---|---|---|---|---|---|
| CDS66501 | Female | Chinese | *LPL* | Hyperlipidemia | Chr8:19955901T>G | NM_000237.3:c.836T>G | p.Leu279Arg | Pathogenic | 851236 |
