## Supplemental Table 6 for "Using HiFi Long-Read Whole Genome Sequencing To Enhance Diagnosis In Patients With Subfertility And/Or Recurrent Pregnancy Loss"

| Individual ID | Gender | Ethnicity | Gene | Disease name | Chromosomal coordinates | NM | HGVS_c | HGVS_p | ACMG Classification | Clinvar ID |
|---|---|---|---|---|---|---|---|---|---|---|
| CDS65401 | Female | Chinese | *IDS* | MPS II (XL) | ChrX:149486091-149494271del | | | | Likely pathogenic | NA |
| CDS67804 | Male | Burmese | *HBA1/2* | Alpha thalassemia | chr16:172240-176044del | NA | -a/-a genotype | NA | Pathogenic | NA |
| CDS69801 | Female | Chinese | *SYNE1* | SCA (AR) | Chr6:152354858G>A | NM_182961.4 | c.10727G>A | p.Arg3576Gln | Conflicting | 402199 |
| CDS69804 | Male | Chinese | *COQ4* | Spastic ataxia (AR) | Chr9:128325849G>A | NM_016035.5 | c.370G>A | p.Gly124Ser | Pathogenic | 476179 |
| CDS70604 | Male | Chinese | *DUOX2* | Thyroid dyshormonogenesis (AR) | chr15:45099686C>A | NM_001363711.2 | c.3391G>T | p.Ala1131Ser | Likely pathogenic | 3577185 |
| | | | *DUOX2* | Thyroid dyshormonogenesis (AR) | chr15:45105775C>T | NM_001363711.2 | c.2202G>A | p.Trp734Ter | Pathogenic | 2736204 |
| CDS72302 | Female | Others | *REN* | Renal tubular dysgenesis | Chr1:204160843-204172821del | | | | Likely pathogenic | NA |
| CDS72303 | Male | Caucasian | *REN* | Renal tubular dysgenesis | Chr1:204160843-204172821del | | | | Likely pathogenic | NA |
| OBG00604 | Male | Malay | *PKHD1* | Polycystic kidney disease (AR) | Chr6:51748297C>T | NM_138694.4 | c.9319C>T | p.Arg3107Ter | Pathogenic | 189090 |
| OBG00704 | Male | Chinese | *ACADSB* | 3 Methylbutyryl coA dehydrogenase def (AR) | Chr10:123037819C>G | NM_001609.4 | c.275C>G | p.Ser92Ter | Pathogenic | 2153596 |
| CHA02 | male | Korean | *ACADVL* | Very long-chain acyl-CoA dehydrogenase deficiency | Chr17: 7222782dup | NM_000018.4 | c.996dup | p.Ala333CysfsTer26 | Likely pathogenic | 370886 |
| CHA03 | female | Korean | *COLQ* | Myasthenic syndrome, congenital, 5 | Chr3:15451658C>T | NM_005677.4 | c.1354C>T | p.Arg452Cys | Conflicting classifications of pathogenicity | 861918 |
| CHA03 | male | Korean | *DNAH5* | Ciliary dyskinesia, primary, 3, with or without situs inversus | Chr5:13753247G>A | NM_001369.3 | c.10858C>T | p.Arg3620Ter | Pathogenic | 454728 |
| CHA04 | female | Korean | *CLN5* | Ceroid lipofuscinosis, neuronal, 5 | Chr13: 77000804TCT>T | NM_006493.4 | c.913_914del | p.Leu305ValfsTer8 | Pathogenic/Likely pathogenic | 1455869 |
| CHA04 | male | Korean | *MCPH1* | Microcephaly, primary autosomal recessive, 1 | Chr8:6499930G>A | NM_024596.5 | c.2214+1G>A | | Likely pathogenic | 2431739 |
| CHA05 | female | Korean | *FANCA* | Fanconi anemia | Chr16:89742841TGTTTT>T | NM_000135.4 | c.3720_3724del | p.Glu1240AspfsTer36 | Pathogenic | 3448 |
| CHA06 | female | Korean | *CYP1B1* | Glaucoma 3, primary congenital, A | Chr2:38071264C>T | NM_000104.4 | c.1090G>A | p.Val364Met | Pathogenic | 1339668 |
| CHA06 | male | Korean | *NMNAT1* | Leber congenital amaurosis 9 | Chr1:9982570C>T | NM_022787.4 | c.709C>T | p.Arg237Cys | Pathogenic/Likely pathogenic | 845745 |
| | | | *NPHS2* | Nephrotic syndrome, type 2 | Chr1:179559710G>A | NM_014625.4 | c.503G>A | p.Arg168His | Pathogenic/Likely pathogenic | 188730 |
| | | | *ALMS1* | Alstrom syndrome | Chr2:73451979C>T | NM_001378454.1 | c.5452C>T | p.Arg1818Ter | Pathogenic/Likely pathogenic | 866994 |
| | | | *ACAD9* | Mitochondrial complex I deficiency, nuclear type 20 | Chr3:128910051C>T | NM_014049.5 | c.1594C>T | p.Arg532Trp | Pathogenic/Likely pathogenic | 30884 |
| 002M | Male | Thai | *GJB2* | Deafness | chr13:20189473C>T | NM_004004.6 | c.109G>A | p.Val37Ile | Pathogenic | 17023 |
| 003F | Female | Thai | *GJB2* | Deafness | chr13:20189473C>T | NM_004004.6 | c.109G>A | p.Val37Ile | Pathogenic | 17023 |
| 004M | Male | Chinese | *F11* | Factor XI deficiency | chr4:186286490G>C | NM_000128.4 | c.1556G>C | p.Trp519Ser | Likely pathogenic | NA |
| 005M | Male | Thai | *JAG2* | Muscular dystrophy, limb-girdle, autosomal recessive 27 | chr14:105145808A>AG | NM_002226.5 | c.2874dup | p.Cys959LeufsTer11 | Likely pathogenic | NA |
| 008F | Female | Thai | *MPO* | Myeloperoxidase deficiency | chr17:58271880C>T | NM_000250.2 | c.1805G>A | p.Trp602Ter | Likely pathogenic | NA |
| | | | *TEK* | Glaucoma 3, primary congenital, E, Venous malformations, multiple cutaneous and mucosal | chr9:27173265C>A | NM_000459.5 | c.804C>A | p.Cys268Ter | Likely pathogenic | NA |
| | | | *G6PD* | Anemia, congenital, nonspherocytic hemolytic, 1, G6PD deficient | chrX-154532269 C>A | NM_001360016.2 | c.1376G>T | p.Arg459Leu | Likely pathogenic | 1722642 |
| 008M | Male | Thai | *F11* | Factor XI deficiency | chr4:186280552C>A | NM_000128.4 | c.1107C>A | p.Tyr369Ter | Pathogenic | 189115 |
| 009M | Male | Thai | *OBSCN* | Rhabdomyolysis, susceptibility to, 1 | chr1:228306469A>AGT | NM_001386125.1 | c.14630_14631dup | p.Val4878TrpfsTer83 | Likely pathogenic | NA |
| 010F | Female | Thai | *KIF12* | Cholestasis, progressive familial intrahepatic, 8 | chr9:114092541C>T | NM_001388308.1 | c.1697+1G>A | p. ? | Likely pathogenic | NA |
| | | | *CEBPE* | Specific granule deficiency | chr14:23118917CAA>C | NM_001805.4 | c.173_174del | p.Phe58CysfsTer7 | Likely pathogenic | NA |
| 010M | Male | Chinese | *ACADM* | Acyl-CoA dehydrogenase, medium chain, deficiency of | chr1:75749418G>A | NM_000016.6 | c.709-1G>A | p. ? | Pathogenic | 370633 |
| | | | *FMO3* | Trimethylaminuria | chr1:171108182CTG>C | NM_001002294.3 | c.591_592del | p.Cys197Ter | Likely pathogenic | NA |
| | | | *F11* | Factor XI deficiency | chr4:186276373G>A | NM_000128.4 | c.738G>A | p.Trp246Ter | Pathogenic | 1457590 |
